# Descriptive Epidemiology of SARS-CoV-2 Gamma (P.1/501Y.V3) variant cases in England, August 2021

**DOI:** 10.1101/2022.05.31.22275827

**Authors:** Nurin Abdul Aziz, Katherine A Twohig, Mary Sinnathamby, Asad Zaidi, Shirin Aliabadi, Natalie Groves, Sophie Nash, Simon Thelwall, Gavin Dabrera

## Abstract

**Purpose:** The Gamma variant of SARS-CoV-2, first detected in travellers from Brazil, was found to have high transmissibility and virulence; following this finding, this paper aims to describe the epidemiology of Gamma cases in England from its first detection on 12 February 2021 to 31 August 2021.

**Methods:** The demographic analysis of Gamma cases was stratified by travel exposure. Travel-associated cases were further analysed by countries travelled from, stratified by categories set in place by the Red (highest risk countries), Amber, Green (lowest risk countries) travel policy, which was implemented from May to October 2021.

**Results:** There were 251 confirmed Gamma cases detected in England in the study period. 35.1% were imported, 5.6% were secondary, and 29.5% were not travel associated. Early cases were predominantly travel-associated, with later cases likely obtained through community transmission. 51.0% of travel-related cases were travellers from Amber countries, and 40.2% had at least one Red country in their journey.

**Conclusion:** The Gamma variant has not seen the same expansion as other variants such as Delta, most likely due to Delta out-competing community transmission of Gamma. Findings indicate the travel policy requiring quarantine for Red and Amber list travellers may have also contributed to preventing onward transmission of Gamma.

## Introduction

The Gamma (P.1/501Y.V3) variant of the Severe acute respiratory syndrome coronavirus 2 (SARS-CoV-2) was first detected through whole-genome sequencing (WGS) in Japan in early January 2021 among a group of travellers arriving from Brazil (1). This variant has since been reported in more than 50 countries (2), including England where the first confirmed case was identified on 12 February 2021.

Evidence suggests that Gamma may be associated with higher transmissibility and propensity for re-infection (3) (4) (5) as well as a possible increased risk of hospitalisation (6); considering the potential increased risks, further investigation into the epidemiology of the variant is necessary. This paper describes the epidemiology of the Gamma variant in England up to 31 August 2021.

## Methods and Data Sources

Individuals infected with the Gamma variant were identified through genotyping by polymerase chain reaction (PCR) and WGS from the national COVID-19 Genomics UK Consortium (COG-UK) sequencing initiative (7) (8). These data were linked to demographic information held in UK Health Security Agency’s Second Generation Surveillance System (SGSS) (9).

Travel exposure was derived from Passenger Location Forms (PLFs), required for entry to the UK, and routine NHS Test and Trace (T&T) surveys. For cases with no known travel from these sources, information came from additional follow up by UKHSA’s Health Protection Teams (HPTs).

Imported cases were defined as confirmed Gamma cases with international travel with arrival into England within 14 days before symptom onset or specimen date. Secondary cases were those who had contact with a PCR-confirmed SARS-CoV-2 case who travelled within 14 days of symptom onset or specimen date. Sporadic Gamma cases had no history of travel nor contact with a SARS-CoV-2 infected traveller. Only T&T and HPT surveys could confirm secondary and sporadic cases. Cases who did not have confirmation of travel were considered unlikely travellers and were excluded from further analysis (n=75, 29.9% of identified Gamma cases).

Descriptive analysis was stratified by travel exposure, with imported and secondary cases grouped as travel-related, examined by demographic factors including, age, sex, region, and place of residence. Place of residence at the time of testing was identified by test location information or unique property registration number (UPRN); the latter was also used to derive data on clustering (10).

For travel-related cases, the associated countries were assessed against the Red, Amber, Green (RAG) categories allocated by the United Kingdom government between May and October 2021, which required hotel quarantine in Managed Quarantine Facilities (MQF) upon return from Red-listed countries and at-home quarantine upon return from Amber-listed countries (12). The RAG assignments were based on the maximum rating of the countries involved in a traveller’s journey at the case’s time of travel. Where RAG assignments were unavailable, an assignment of Red was given to those who travelled from a country on the initial travel ban list of countries associated with the Gamma variant (11).

## Findings

Between 12 February and 31 August 2021, there were 251 confirmed Gamma cases detected in England. Of these, 88 (35.1%) were imported cases, 14 (5.6%) were secondary cases associated with travel, and 74 (29.5%) were sporadic. Early Gamma cases were mostly imported or secondary cases; however, cases since the 12^th^ April were mainly sporadic (Figure 1). There were no reported deaths among confirmed Gamma cases in England during the study period.

**Figure 1.**
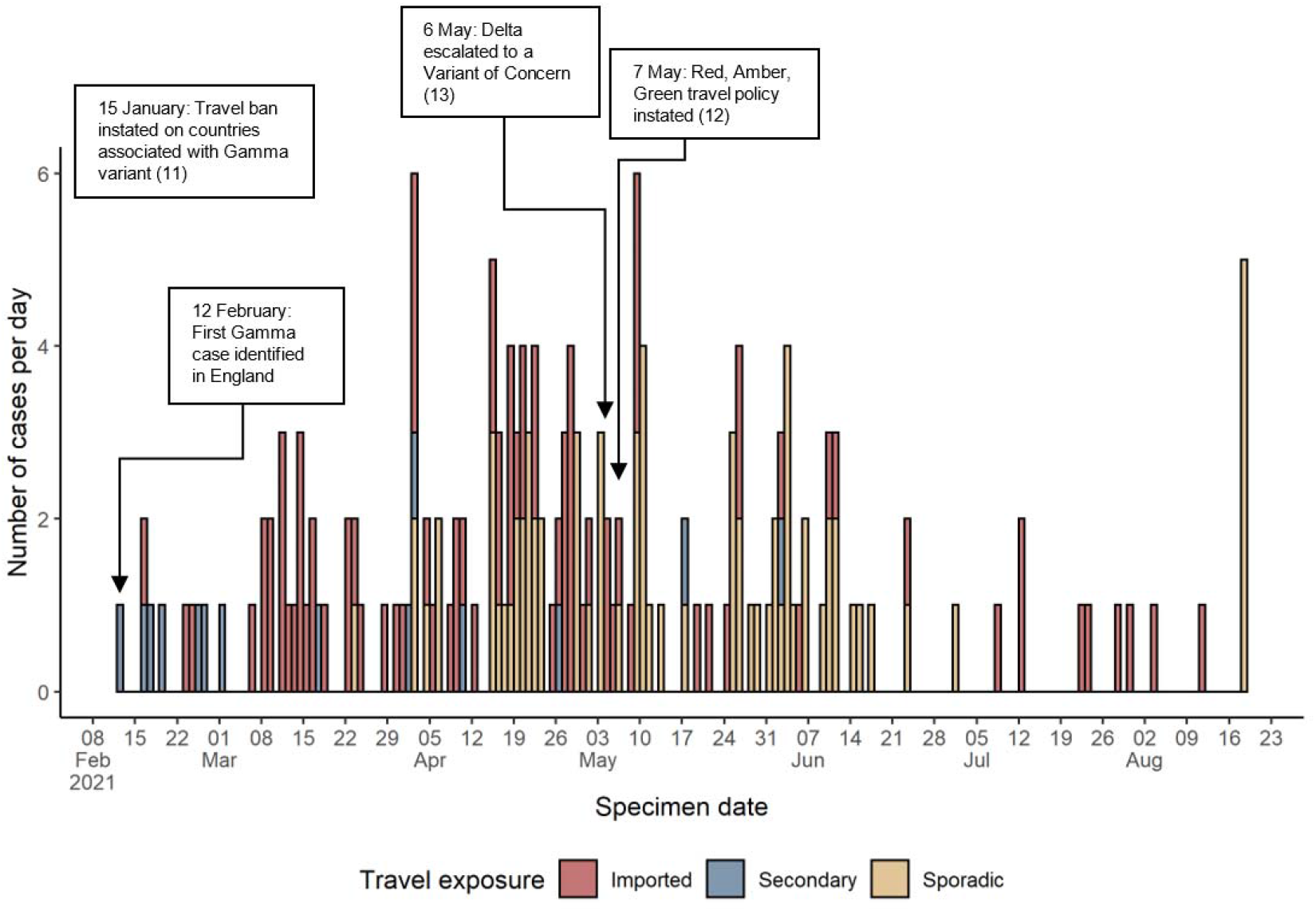
Epidemic curve of SARS-CoV-2 Gamma (P.1) cases in England by travel exposure, from 12 February 2021 to 31 August 2021, including key events. Gamma cases with unknown travel exposure were omitted.

### Demographic distribution

The age group with the highest number of cases for travel-related (n=33, 32.4%) and sporadic cases (n=19, 25.7%) was the 20-29 group. About half of the cases for both travel-related and sporadic cases were from the London region.

### Travel-related cases

Half of travel-related cases (n=52, 51.0%) were travellers to at least one Amber country as the highest rating; 40.2% (n=41) had at least one Red country in their journey. Among all travel-related cases, the highest number of cases reported travel from Brazil (n=27, 21.4%), which was on the Red list since its first publication in May 2021, followed by Mexico (n=15, 11.9%), an Amber country upgraded to Red on 8 August 2021, and Amber countries USA (n=14, 11.9%) and France (n=12, 11.1%).

### Place of residence and clusters

There were 25 cases associated with MQFs, however most travel-related (n=56, 54.9%) and sporadic cases (n=68, 91.9%) were reported from residential dwellings at the time of testing.

Most travel-related cases were not part of a residential cluster (n=74, 72.5%), whereas a larger proportion of sporadic cases were part of a residential cluster (n=31, 41.9%), with cluster size ranging from 2 to 6.

## Discussion & conclusions

This analysis shows that earlier cases of the Gamma variant in England were predominantly travel-associated, transitioning to more cases likely obtained through community transmission. However, the Gamma variant has not seen the same expansion as other variants such as Delta (14), despite initial concerns of increased virulence and transmissibility. It is likely that Delta was able to out-compete community transmission of Gamma; while this was not previously analysed in England, Delta was observed to have surpassed Gamma in Brazil and Mexico (15) (16).

However, there is a bias in the detection of travel-associated cases due to targeted sequencing of new arrivals and their contacts and routine testing of travellers (17). Community-transmitted cases are therefore more likely to be underestimated, as they are subject to randomised population-level sample-picking processes rather than targeted sequencing; however, sequencing coverage during the peak of incidence of Gamma between March and June remained above 50% (14), indicating the likelihood that the majority of community cases were identified.

The RAG policy changed in October 2021 such that countries are no longer placed on an Amber or Green list (12); hence, quarantine is currently only required for Red-list travellers. The analysis indicates the policy requiring quarantine in MQFs for Red-list travellers and at-home quarantine for Amber-list travellers, implemented during the study period, may have contributed to preventing onward transmission. The effect of this is shown in the small numbers of secondary cases, and that most travel-related cases were not linked to residential clusters of other COVID-19 cases.

It should be noted that as of April 2022, routine surveillance of travellers into England has ceased: PLFs ceased being mandatory on 18 March 2022 (17), and contact tracing by T&T ended on 24 February 2022 (18). This study supports that surveillance related to travel from high and medium risk countries was important to inform efforts to control travel-related Gamma infections. While larger scale surveillance of travellers has ceased, a similar approach should also be considered when new variants emerge, such as using existing surveillance systems such as surveys implemented by local HPTs to collect data on the travel status of variant cases.

## Data Availability

This analysis was based on routine individual-level healthcare data, which cannot be made available to others by the study authors.

## Authors’ approval and contributions

All authors contributed to the study conception and design. Data collection and preparation were performed by NA, KT, MS, AZ, SA, NG, and SN. Data analysis was performed by NA. The first draft of the manuscript was written by NA, and all authors commented on previous versions of the manuscript. All authors read and approved the final manuscript.

## Tables

**Table 1.** Demographic description of SARS-CoV-2 Gamma (P.1) cases in England. Data up to 31 August 2021.

|  | Imported/Secondary (n = 102) |  | Sporadic (n = 74) |  |
| --- | --- | --- | --- | --- |
| Age | n | % | n | % |
| <10 | 4 | 3.9 | 6 | 8.1 |
| 10-19 | 8 | 7.8 | 15 | 20.3 |
| 20-29 | 33 | 32.4 | 19 | 25.7 |
| 30-39 | 31 | 30.4 | 15 | 20.3 |
| 40-49 | 12 | 11.8 | 9 | 12.2 |
| 50-59 | 9 | 8.8 | 8 | 10.8 |
| 60-69 | 2 | 2.0 | 2 | 2.7 |
| 70-79 | 3 | 2.9 | 6 | 8.1 |
| 80+ | 4 | 3.9 | 15 | 20.3 |
| **Sex** |  |  |  |  |
| Female | 44 | 43.1 | 37 | 50.0 |
| Male | 58 | 56.9 | 36 | 48.6 |
| Unknown | 0 | 0.0 | 1 | 1.4 |
| **Region<sup>a</sup>** |  |  |  |  |
| London | 57 | 55.9 | 39 | 52.7 |
| Midlands and East of England | 15 | 14.7 | 14 | 18.9 |
| North of England | 4 | 3.9 | 10 | 13.5 |
| South of England | 21 | 20.6 | 9 | 12.2 |
| Unknown | 5 | 4.9 | 2 | 2.7 |
| **Property classification** |  |  |  |  |
| Residential dwelling (inc. houses, flats) | 56 | 54.9 | 68 | 91.9 |
| Managed quarantine facilities | 25 | 24.5 | 0 | 0.0 |
| Other property classifications | 4 | 3.9 | 2 | 2.7 |
| Undetermined | 17 | 16.7 | 4 | 5.4 |
| **RAG assignment<sup>b</sup>** |  |  |  |  |
| Red | 41 | 40.2 | - | - |
| Amber | 52 | 51.0 | - | - |
| Green | 1 | 1.0 | - | - |
| Unknown | 8 | 7.8 | - | - |

<sup>a</sup> The nine regions of England were aggregated to form four larger regions to suppress small numbers and facilitate comparison: North of England (North West, North East England), Midlands and East of England (East Midlands, West Midlands, East of England), South of England (South West, South East England), and the London region.
<sup>b</sup> Red Amber Green (RAG) assignments based on the country with the most restrictive travel rules in a traveller's journey (i.e. Red > Amber > Green); this includes journeys of imported variant cases and journeys of travellers in contact with secondary cases.

## References

1. National Institute of Infectious Diseases, Japan. Brief report: New Variant Strain of SARS-CoV-2 Identified in Travelers from Brazil. Japan : National Institute of Infectious Diseases, Japan, 2021.

2. PANGO Lineages. Lineage P.1. PANGO Lineages. [Online] 22 January 2021. [Cited: 28 June 2021.] https://cov-lineages.org/global_report_P.1.html.

3. Genomics and epidemiology of the P.1 SARS-CoV-2 lineage in Manaus, Brazil. Faria, Nuno R, et al. 6544, s.l. : Science, 21 May 2021, Vol. 372, pp. 815–821.

4. Naveca, Felipe, et al. COVID-19 epidemic in the Brazilian state of Amazonas was driven by long-term persistence of endemic SARS-CoV-2 lineages and the recent emergence of the new Variant of Concern P.1. [Preprint]. s.l. : Research Square, 25 February 2021.

5. Public Health England. SARS-CoV-2 variants of concern and variants under investigation in England - Technical briefing 07. s.l. : Public Health England, 2021. Technical briefing.

6. Characteristics of SARS-CoV-2 variants of concern B.1.1.7, B.1.351 or P.1: data from seven EU/EEA countries, weeks 38/2020 to 10/2021. Funk, Tjede, et al. 16, 22 April 2021, Euro Surveillance, Vol. 26.

7. Bull, Matt, et al. Standardised Variant Definitions. [Online] 29 Abdel Latif, Alaa, et al. Mexico Mutation Report. outbreak.info. [Online] https://outbreak.info/location-reports?loc=MEX.

8. Department of Health and Social Care. Reflex (genotyping) assays for identification of priority SARS-CoV-2 variants of concern. 2021.

9. Timeliness and completeness of laboratory-based surveillance of COVID-19 cases in England. Clare, Tom, et al. May 2021, Public Health, Vol. Volume 194, pp. 163–166.

10. Penetration and impact of COVID-19 in long term care facilities in England: population surveillance study. Chudasama, Dimple Y, et al. 1 September 2021, International Journal of Epidemiology.

11. Department for Transport; Public Health England. Travel from South American destinations, Portugal, Panama and Cape Verde banned to prevent spread of new variant. [Online] 2021. https://www.gov.uk/government/news/travel-from-south-american-destinations-portugal-panama-and-cape-verde-banned-to-prevent-spread-of-new-variant.

12. Department of Transport; Department of Health and Social Care; Public Health England. Red, amber, green lists: check the rules for travel to England from abroad. [Online] 7 May 2021. Page now archived after new travel guidance released on October 2021. https://webarchive.nationalarchives.gov.uk/ukgwa/20210831163431/https://www.gov.uk/guidance/red-amber-and-green-list-rules-for-entering-england.

13. Public Health England. SARS-CoV-2 variants of concern and variants under investigation in England - Technical Briefing 10. s.l. : Public Health England, 2021. Technical briefing.

14. Public Health England. SARS-CoV-2 variants of concern and variants under investigation in England - Technical briefing 18. s.l. : Public Health England, 2021. Technical briefing.

15. Patané, José, et al. SARS-CoV-2 Delta variant of concern in Brazil - multiple introductions, communitary transmission, and early signs of local evolution. [Preprint]. s.l. : MedRxiv, 22 September 2021.

16. Abdel Latif Alaa, et al. Mexico Mutation Report. outbreak.info. [Online] https://outbreak.info/location-reports?loc=MEX.

17. Hospital admission and emergency care attendance risk for SARS-CoV-2 delta (B.1.617.2) compared with alpha (B.1.1.7) variants of concern: a cohort study. Twohig, Katherine A, et al. 27 August 2021, Lancet Infectious Diseases.

